# Factors Associated with the Risk of HIV Infection among HIV-exposed Infants in Malawi: 2013-2020

**DOI:** 10.1101/2021.10.06.21264671

**Authors:** Wingston Ng’ambi, Janne Estill, Fatma Aziza Merzouki, Erol Orel, Tiwonge Chimpandule, Rose Nyirenda, Olivia Keiser

## Abstract

**Background:** Despite the high availability of individual-level data of infants accessing HIV DNA polymerase chain reaction (DNA-PCR) testing service, there has been little in-depth analysis of such data. Therefore, we describe spatial and temporal trends in risk of HIV infection among Malawi’s HIV-exposed infants (HEI) with DNA-PCR HIV test result from 2013 to 2020.

**Methods:** This is an implementation study using routinely collected patient-level HIV DNA-PCR test result data extracted from the national Laboratory Management Information System database managed by the Department of HIV/AIDS between 1 January 2013 and 30 June 2020. We calculated frequencies, proportions and odds ratios (OR) with their associated 95% confidence intervals (95%CI). We performed a random-effects logistic regression to determine the risk factors associated with HIV infection in infants, controlling for the spatial autocorrelation between districts and adjusting for other variables.

**Results:** We evaluated 255,229 HEI across 750 facilities in 28 districts. The overall risk of HIV infection among all tested HEI between 2013 and 2020 was 7.2% (95%CI: 7.1-7.3). We observed a decreasing trend in the proportion of HEI that tested HIV positive from 7.0% (95%CI: 6.6-7.4) in 2013 to 5.7% (95%CI: 5.4-5.9) in 2015 followed by an increase to 9.9% (95%CI: 9.6-10.2) in 2017 and then a decreasing trend to 4.2% (95%CI: 3.7-4.6) in 2020. The risk of HIV infection increased by age of the HEI. There was spatial heterogeneity of HIV prevalence between districts of Malawi.

**Conclusion:** We summarised spatial and temporal trends of risk of HIV infection amongst HEI in Malawi between 2013 and 2020. There is need for further strengthening of EID program to ensure that all the HEI are enrolled in care by eight weeks of age in order to further reduce mother-to-child transmission of HIV.

**Key message:** There is need for further strengthening of the Malawi early infant diagnosis program to ensure that all the HIV-exposed infants are enrolled in care by eight weeks of age in order to eliminate mother-to-child transmission of HIV by 2030.

**What is known about the study?:** Malawi has implemented the Early Infant Diagnosis (EID) since 2009. Most of the studies on EID in Malawi have focused on just one or two health facilities.

**What the study adds?:** This is to our knowledge the first in-depth analysis of national routine data on HIV DNA-PCR tests among HIV-exposed infants with in Malawi. Our study has shown that there is spatial and temporal heterogeneity in risk of HIV infection amongst the HEI in Malawi between 2013 and 2020.

## INTRODUCTION

Despite the tremendous global progress in the HIV response, children continue to be affected substantially by the epidemic [1]. Of the estimated 38.0 million people living with HIV worldwide in 2020, 2.8 million were children aged 0-19 [1]. Globally, most of the children living with HIV are found in Africa. Sub-Saharan Africa has the largest burden of paediatric HIV in the world [1] [2]. In Malawi, the HIV estimates from the Spectrum software indicate approximately 2500 children living with HIV and 1800 AIDS deaths among children aged below fifteen years in 2020.

Although several countries like Malawi, South Africa and Uganda [3] have registered very high uptake of prevention of mother to child transmission services, the uptake of services for HIV-exposed infants have been suboptimal for various reasons in most low and middle income countries (LMIC) [4]. Diagnosis of paediatric HIV has been one of the major challenges in resource-limited settings leading to lower proportion of children living with HIV who start antiretroviral therapy (ART) compared with adults [5]. The WHO guidelines recommend HIV ascertainment for exposed infants (HEI) as part of routine care, as early as 6 weeks of age [5]. In Malawi, HIV DNA polymerase chain reaction (DNA-PCR) is commonly used to test HIV in HEI at registration in the Early Infant Diagnosis (EID) programme [6].

Malawi started the EID programme in 2009 following a recommendation by the WHO so that all infants exposed to HIV during pregnancy, labour, delivery and breastfeeding have HIV status ascertainment by the age of 6 weeks with follow up HIV tests at 12 and 24 months[6]. HIV ascertainment among HEI is critical in facilitating provision of life-saving treatment for those infected with the virus and enables access to HIV prevention information and support for those testing negative. Currently over 650 facilities are providing EID services in Malawi. The Malawi Ministry of Health as well as the PEPFAR supported programmes track HIV prevalence among HIV-exposed infants. Since the introduction of this program no in-depth analyses have been done to assess the trends of HIV prevalence amongst the HEI tested EID DNA PCR at a national level. However, in-depth analyses are necessary for a greater understanding of HIV prevalence for EID program quality improvement.

Furthermore, in-depth analyses of the EID program are necessary in tracking the first and second steps of the UNAIDS 95-95-95 target for ending HIV/AIDS by 2030. This study therefore aims to describe HIV prevalence trends and assess the factors associated with trends in the risk of HIV infection of HEI tested with DNA-PCR in Malawi between 2013 and 2020.

## METHODS

### Study design

This is an implementation study involving a retrospective review of patient-level HIV DNA-PCR data obtained from the National Laboratory Information Management Systems (LIMS) national database containing data collected between 2013 and 2020 in Malawi [7]. The LIMS database is managed by MOH Diagnostics in the Department of Technical and Support Services (HTSS). The LIMS database contains individual level DNA-PCR data for HIV ascertainment amongst the HIV exposed infants aged 24 months and below. Data across all the districts and facilities are included. The database has inbuilt tools for performing data quality assessment like range checks and other associated validation rules. The data are entered at the DNA-PCR laboratories in Malawi. By 30 June 2020, there were 10 laboratories performing DNA-PCR HIV testing for HIV-exposed infants in Malawi: Dream laboratory in Blantyre, Dream Laboratory in Balaka, Kamuzu Central Hospital, Mzimba District Hospital, Mzuzu central Hospital, Nsanje District Hospital, Partners in Hope, Queen Elizabeth Central Hospital, Thyolo District Hospital, and Zomba Central Hospital.

### Management of HIV exposed infants in Malawi

The management of HIV-exposed infants (HEI) is based on the Malawi ART/PMTCT guidelines [6]. The HEI are registered in the Early Infant Diagnosis (EID) Programme at 6 weeks after birth a HIV DNA–PCR test is conducted during the registration into the EID programme. In addition, HEI are put on Cotrimoxazole Preventive Therapy (CPT) to prevent certain opportunistic infections [6]. Rapid HIV diagnostic tests are done at 12 months and 24 months or as necessary [6].

### Statistical Analysis and Data Management

The data were managed in Stata v16.0 (Stata Corp., Texas, USA). The response variable was HIV infection status. The independent variables were: age (in months) at sample collection, year sample collected, sex, facility location (rural/urban) and region (north/centre/south). A descriptive analysis was first performed detailing the characteristics of the study population. We also fitted bivariate analysis of each of the independent variable and HIV status. Only the independent variables that were statistically significant at 20% were eligible for inclusion in the multivariable model. We fitted a multivariable logistic regression model of HIV infection using a forward step-wise selection method, with age and sex entered as a priori variables.

Since HIV prevalence varies by district, we controlled for random clustering effect of the district when conducting logistic regression of the independent variables on HIV infection. We presented both crude and adjusted odds ratios (OR) of HIV infection of each independent variable. Multiple imputation chained equations (MICE), with five imputation rounds and 5000 permutations, were used to impute missing data of the following covariables: age category when sample was taken, HIV status, child’s sex and year of sample collection. The analysis produced the within district variation (rho) and between district variation (σ) of the risk of HIV infection due to controlling for clustering effect of the district. We resented the annual the risk of HIV infection for all the districts of Malawi using a forest plot of the pooled the risk of HIV infection by districts in order to get the degree of heterogeneity of the risk of HIV infection by districts. Statistical significance was set at P<0.05.

### Patient and public involvement statement

Over the recent years in the implementation of HIV EID programme in Malawi, there has been need to test the HIV status of the HIV-exposed infants to determine their risk of HIV infection. This risk ascertainment begins with the enrolment of HEI into the EID programme. Every mother of HEI undergoes a counselling session in order to be sensitized on the follow-up of her child in the EID programme. The mothers of HEI ensures that the HEI get enrolled and followed up in the EID programme. All follow-up processes conform to the national HIV treatment guidelines. The results of this study will be shared with the HIV programme managers across health facilities of Malawi. This will ensure that the results inform practice at both facility and national levels.

### Ethical approval

The study was approved by the Malawi National Health Sciences Research Committee (NHSRC) in Lilongwe, Malawi (protocol #: 1669). As this study used secondary anonymised data, no informed consent was needed.

## RESULTS

### Characteristics of HIV-exposed infants who had DNA-PCR HIV test

The characteristics of HIV exposed infants (HEI) with HIV DNA-PCR testing are shown in Table 1. We evaluated 255,229 HIV exposed infants with DNA-PCR results. Of these, 145,622 (57%) had HIV DNA-PCR testing done before two months after birth. The numbers of males and females tested for HIV were similar, 159,699 (63%) were from the southern region while 22,897 (9%) were from the northern region (Table 1). We observed an increasing trend in the number of HEI tested for HIV from 16,308 (6%) in 2013 to 43,370 (17.0%) in 2018 and a decrease thereafter (Table 1). The proportion of missing data ranged from 2.9% (7,344 of 255,229) for sex of the child to 3.3% (8,354 of 255,229) for age at sample draw.

**Table 1:**
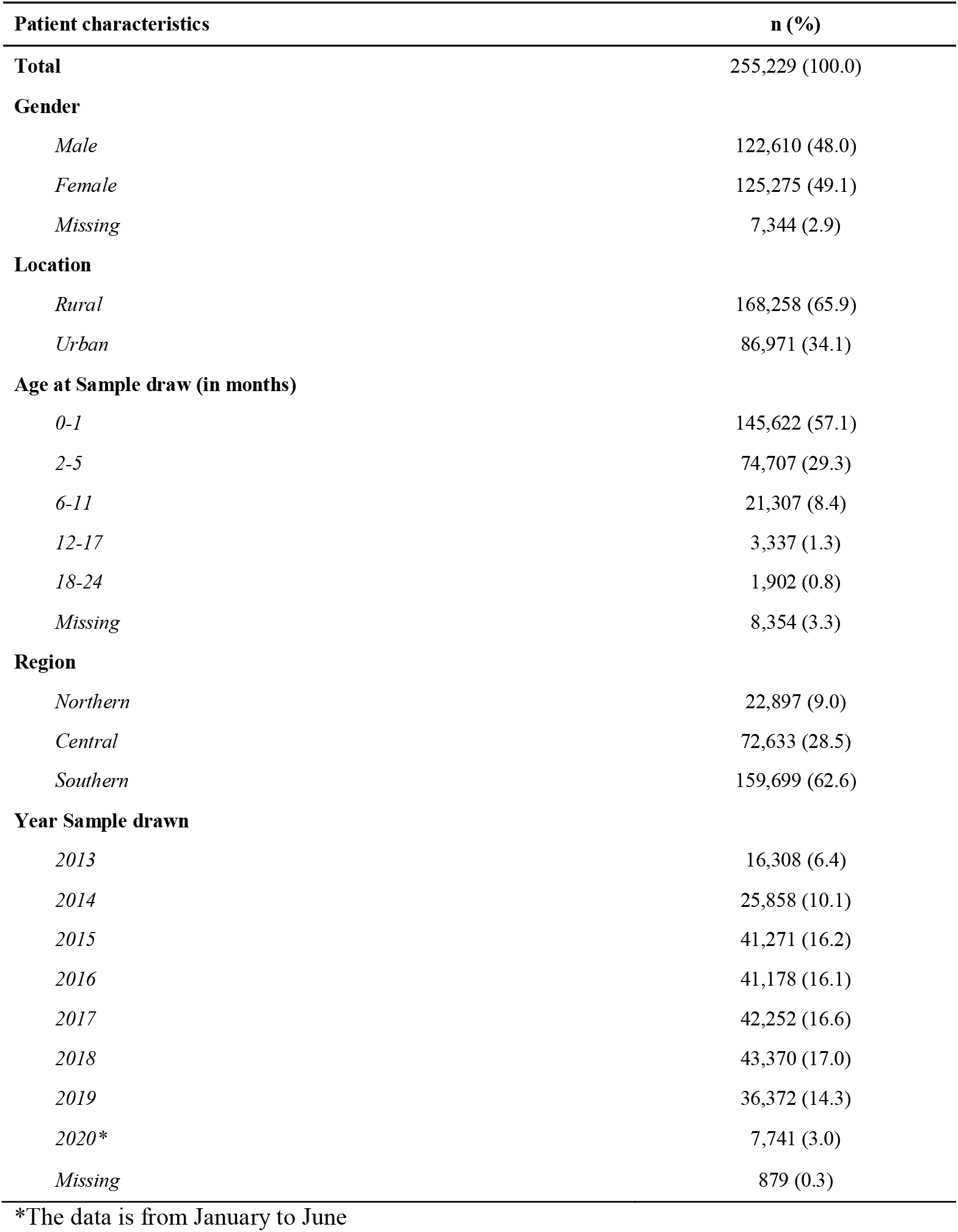
Characteristics of HIV-exposed Infants with DNA-PCR HIV test results between 2013 and 2020 in Malawi.

### Risk of HIV infection among HIV-exposed infants with HIV DNA PCR HIV test

A total of 235,774 (92%) children had complete data on location and region as shown in Table 2. We observed that 16,936 (7.2%. 95%CI: 7.1-7.3) of the 235,774 HEI had positive HIV DNA-PCR results. The female and male HEI had similar HIV infection risk (see Table 2). There was an increasing trend in the risk of HIV infection with age at HIV testing. The southern region had the lowest risk of HIV infection among HEI while the northern region had the highest risk of HIV infection.

**Table 2:**
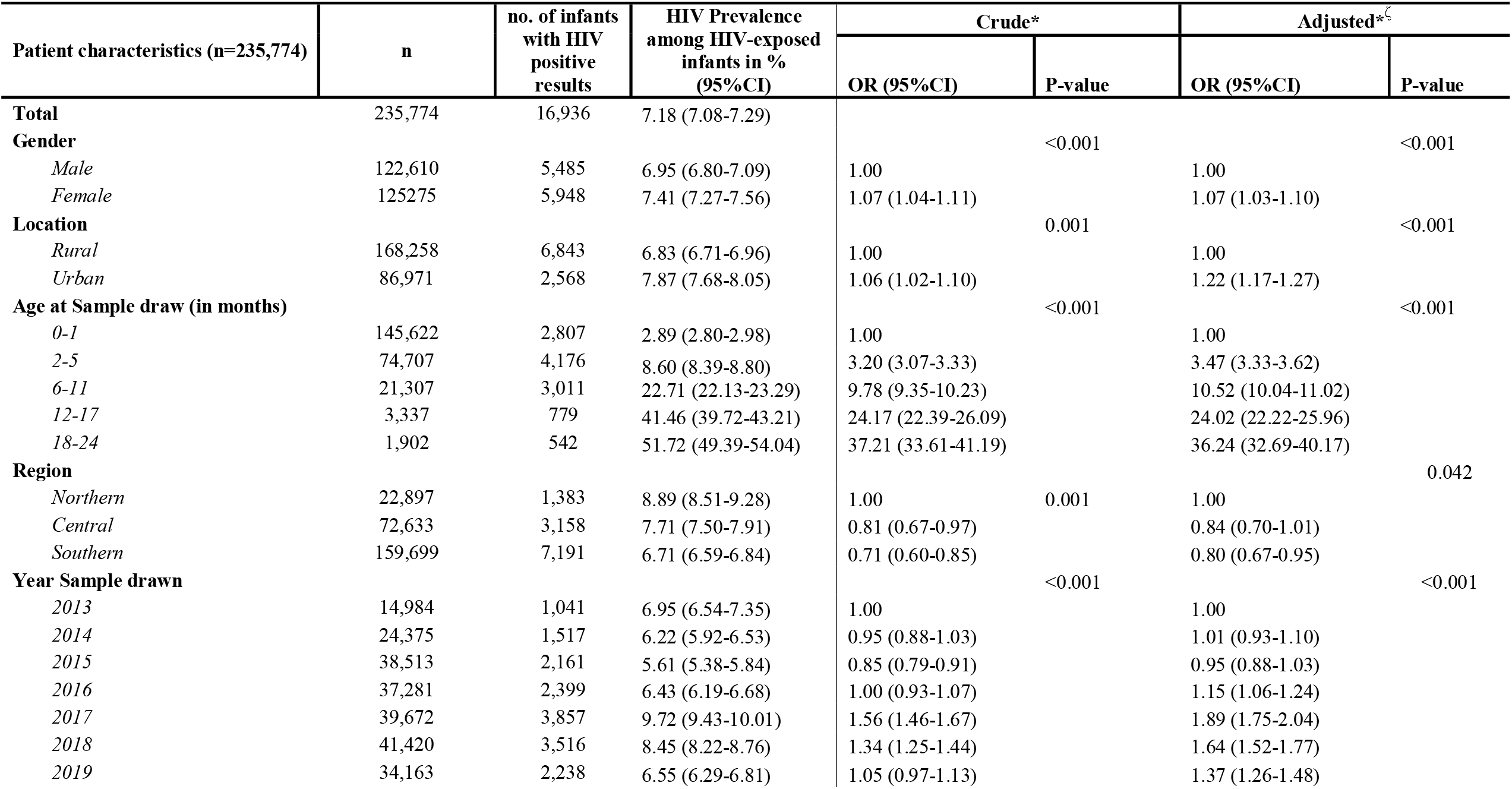
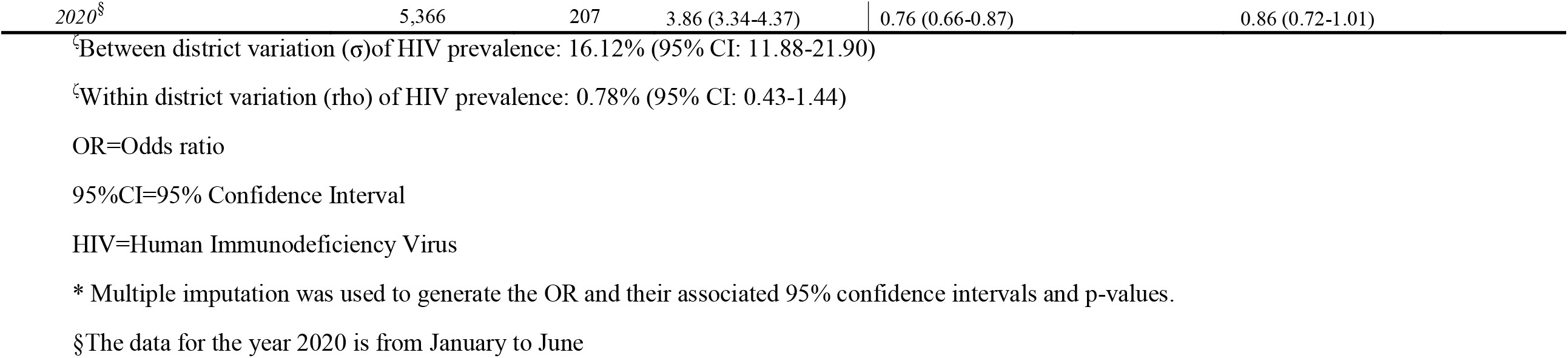
Factors associated with HIV prevalence among HIV-exposed infants with DNA-PCR HIV tests between 2013 and 2020 in Malawi.

### Temporal distribution of the HIV infection risk

The trend in risk of HIV infection across the regions is shown in Figure 1. The overall risk of HIV infection dropped from 7.0% (95%CI: 6.6-7.4) in 2013 to 5.7% (95%CI: 5.4-5.9) in 2015 followed by an increase to 9.9% (95%CI: 9.6-10.2) in 2017 and then a decreasing trend to 4.2% (95%CI: 3.71-4.63) in 2020. Between 2015 and 2017, the northern, central and southern regions experienced an increase in the trend of risk of HIV infection (see Figure 1). There was strong evidence of association between the risk of HIV infection and region of residence of the HEI in Malawi between 2013 and 2020.

**Figure 1:**
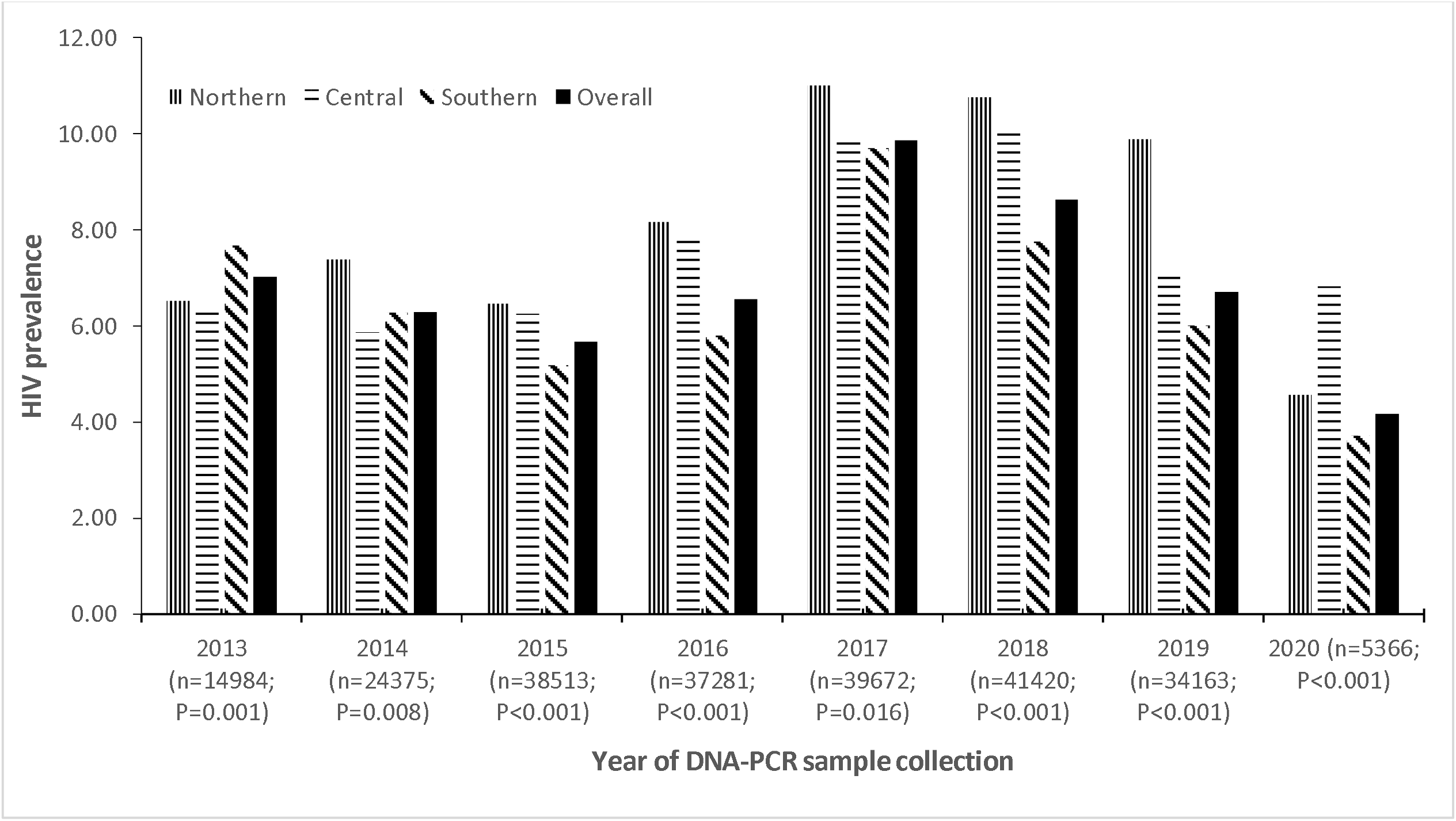
Temporal Trend in HIV prevalence amongst HIV exposed infants in Malawi across the Northern, Central and Southern Regions between 2013 and 2020. DNA-PCR= HIV DNA Polymerase Chain Reaction Tests

### Factors associated with the risk of HIV infection

The factors associated with the risk of HIV infection are shown in Table 2. The adjusted odds of HIV infection among female HEI were 1.07 (95%CI:1.03-1.10, P<0.001) times those of male HEI. There was increasing odds of HIV infection by age at HIV testing (AOR=3.47; 95%CI: 3.33-3.62 and AOR=36.24; 95%CI:32.69-40.17) amongst those aged 2-5 and 18-24 months respectively compared to those aged less than 2 months at HIV DNA-PCR sample collection). Infants residing in urban areas had higher odds of HIV infection compared to those living in rural areas. After adjusting for age, sex, location and region, the infants that were tested between 2016 and 2019 were more likely to be HIV positive compared to those tested in 2013 (see Table 2).

### Spatial distribution of HIV infection

The spatial distribution of HIV amongst the HIV-exposed infants is shown in Figure 1. There was a strong association between HIV infection and district of residence (P<0.01). Within each district, the risk of HIV infection varied by 0.78% (95%CI: 0.42-1.44) over the 2013-2020-time period as shown in Table 2. However, there was variation in risk of HIV infection across the districts (σ=16.13%; 95%CI: 11.88-21.90; P<0.001) as shown in Table 2. Some districts had the risk of HIV infection as high as 9.9% while in others it was as low as 4.6% between 2013 and 2020 as shown in Figure 2. The six districts with the highest risk of HIV infection among HEI were Lilongwe, Likoma, Nkhotakota, Chitipa, Karonga and Nkhata Bay while the lowest risk of HIV infection was observed in Neno, Chiradzulu, Phalombe, Thyolo, Mulanje and Dedza.

**Figure 2:**
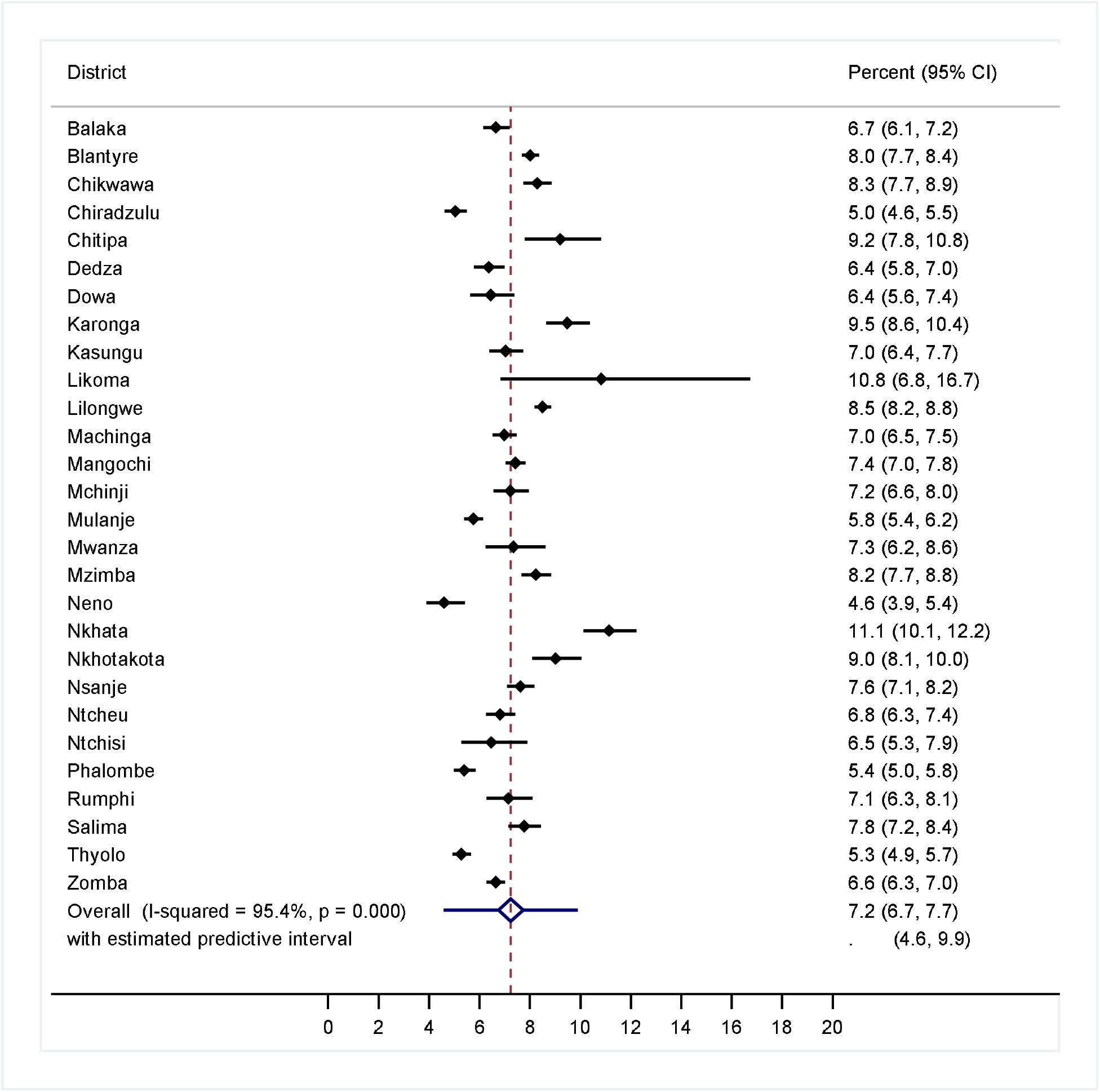
Pooled HIV Prevalence among the HIV Exposed Infants with HIV-DNA PCR Tests in Malawi between 2013 and 2020.

## DISCUSSION

This is a national analysis of HIV DNA-PCR data obtained from the laboratory management information system (LIMS) in Malawi. The overall risk of HIV infection amongst the DNA-PCR tests was high implying the need to strengthen the MTCT programme. We observed increasing trend in probability of HIV infection by age at sample collection. The highest risk of HIV infection was observed among the HEI tested in 2017. HIV infection differed across the districts of origin of the HEI, and the highest risk of HIV infection were observed amongst the HEI from the Northern region. Furthermore, the odds of HIV infection were higher in urban than in rural areas.

The risk of HIV acquisition among infants exposed to HIV in our study was almost two times higher than that observed in South Africa [8] [9] but similar to other studies conducted in Malawi and settings India [10] [11] [16]. Consistent with other studies, the risk of HIV infection of the HEI was higher with older age at DNA-PCR sample collection [12]. With the high risk of HIV infection being observed among EID in the Northern Region of Malawi, it is imperative to consider the northern region with quality improvement projects aimed at bringing down the HIV infection risk among HEI. The Northern Region does not have as many HIV implementing partners as the other regions due to funding prioritization; among the general population HIV infection risk is substantially higher in the southern and central regions than the northern region [13]. Although the risk of HIV infection has been reported to be higher amongst the female than the female population [13], we observed similar risk in HIV infection by sex of the child.

Our findings also demonstrate considerable heterogeneity in risk of HIV infection among the HEI in Malawi. Several spatial epidemiological studies indicate spatial variation of diseases which could be attributed to social and cultural factors [14] [15] [16]. Generally, studies of HIV epidemiology in Malawi have been highly predominant in the districts in the southern region followed by the central and northern regions [13] [17]. The spatial pattern of the risk of HIV infection would imply the need to target PMTCT interventions in the districts with high risk of paediatric HIV acquisition in order to improve the health of the children and the women [13].

There has been a temporal trend in HIV infection risk by year. This is consistent with many studies and surveys conducted in Malawi that have shown a downward trend in the risk of HIV infection [13]. The upward increase in the risk of HIV infection of the HIV exposed infants may have occurred as a result of a weaker implementation of PMTCT services especially with regard to follow-up of HEI which has been reported in Malawi [10]. This is also consistent with what has been observed in Sub-Saharan African settings with the general decreasing trend in HIV infection among the HIV exposed infants. Possible explanations to the downward trend in HIV infections include successful implementation of PMTCT programmes and the high antiretroviral therapy coverage in general [18] [19].

The major strength of this study is the large sample size and being conducted within a routine programme setting, which has the potential to improve the EID programmes in Malawi and similar settings. The major limitation of this study is that the data in LIMS only cover baseline data with no follow-up tests conducted with rapid HIV diagnostic tests at 12 and 24 months. The Department of HIV/AIDS of the Malawi ministry of Health and Population should make an attempt to have all HIV laboratory tests of the HIV-exposed infants recorded in the LIMS. Such data should be managed in a way that it would be possible to track the HIV-exposed infant throughout the 24 months of follow-up in the EID programme. Another limitation is that the data are not linked to data on ART initiation among the HIV-exposed infants that were found to be HIV positive. Furthermore, the maternal information was not captured in LIMS hence we could not include such information in this analysis. Having such data would provide more information on the risk factors for HIV infection on HEI.

## CONCLUSION

In conclusion, this is to our knowledge the first in-depth analysis of national routine data on HIV DNA-PCR tests among HIV-exposed infants with in Malawi. Our study has shown that there is spatial and temporal heterogeneity in risk of HIV infection amongst the HEI in Malawi between 2013 and 2020. There is a need for further strengthening the EID program to ensure that all the HEI are enrolled in care by eight weeks of age. As this study only looks at HIV DNA-PCR test results, there is a need for a follow-up study examining risk of HIV infection in the entire twenty-four months of follow-up in order not to underestimate or over-estimate the true risk of HIV infection. Access to HIV DNA-PCR testing will ensure that 90% of the HEI with HIV will have known HIV status hence supporting the way towards reaching the 95-95-95 target HIV strategy by 2030 in Malawi [19].

## Data Availability

Data should be requested from the corresponding author

## DECLARATIONS

We declare that there is no conflict of interest in publishing this paper.

## AUTHORS’ CONTRIBUTIONS

WN led the manuscript writing, conducted data management and analysis; FAM advised on the data analysis and policy insights on the paper; JE advised on data analysis and policy insights on the paper; EO advised on data analysis and policy insights on the paper and OK advised on the analysis and policy insights on the paper. All authors read and approved the final manuscript.

## ACKNOWLEDGEMENTS

The study was supported by a grant from the Swiss National Science Foundation (no 163878). The authors would also like to thank all the parents of the HIV exposed infants together with the HEI that participated in this study. Further, the authors thank all the health facility staff that supported DNA-PCR sample collection and processing as well as counselling of the mothers of the HEI. The authors would also like to thank the Department of HIV/AIDS for allowing them to extract and analyse the data.

